# Polygenic Risk Score Comparative Analyses Reveals Risk Disparity of Genetic Predisposition to Chronic Kidney Disease- A Multi Ancestry Approach

**DOI:** 10.1101/2022.07.05.22277245

**Authors:** Varun Sharma, Indu Sharma, Love Gupta, Garima Rastogi, Anuka Sharma

## Abstract

Polygenic Risk Score (PRS) models are used extensively to find the population/individual risk towards disease. These predictive scores are of great help as risk scores if predicted earlier the life of individual can be saved from the chronic/ complex diseases. In this empirical assessments study, the polygenic risk score was calculated in three different ancestries (SAS, EAS and African Americans) based on more than three hundred markers. The risk score we observed indicated that average population risk scores are varied but on cumulating the ancestries the average risk score increased ∼1.3 times than individual population average risk. The parameter which varies greatly while calculating the PRS is the ancestry; it should be prerequisite that individuals of same ancestry should be taken as a one population groups while calculating the scores.

## Introduction

Chronic Kidney Disease (CKD) is a progressive disease defined by glomerular filtration rate lesser than 60ml/min/1.73m^2^along with other medical conditions if these persisting more than three months like albuminuria should be 30mg per 24 hours or polycystic/ dysplastic kidneys or hematuria[1].On an average 10-16% of the general population is affected by CKD and is having high mortality and morbidity as being unrecognized by patients and even by clinicians [2-4] and has become a major public health issue. Global all age prevalence of CKD has increased from 29.3% to 41.5% between the years 1990 to 2017 [5].

Diabetes Mellitus (DM) or hypertension are the hallmarks for CKD globally, its prevalence is higher in developing countries with additional factors to DM and CKD such as glomerulonephritis, infection and huge exposure to air pollution, pesticides and herbal remedies used [4]. As per the pathogenic succession of kidney disease, patient having CKD are at higher risk for developing end stage renal disease (ESRD). Kidney dysfunctions can be observed with increased levels serum levels of cystatin C, creatinine or urea. The best marker studied till date for CKD is GFR which is measured using exogenous markers or estimated (eGFR) depending on the concentrations of endogenous filtration markers of serum creatinine and cystatin C[6, 7]. For the longer survival of the individual diagnosed with ESRD requires dialysis or a kidney transplant to maintain the survival of the patient [8].

With the increase in the percentage of the disease numerous GWAS (Genome Wide Association Studies) have been conducted and many variants are identified in the last decade for CKD[7, 9-12]. This has also leaded to increase in testing out the different models of PRS (Polygenic Risk Score) for kidney risks in individual and in population cohort [13-15]. PRS is integration of mathematical aggregation of risk derived from the variants on the DNA present across the genome [16]. This score will help in knowing the risk factors in advance which might be higher in the coming years in population set or in an individual. The group/individual which is at higher risk as per the score can be highly benefited in controlling the disease with better treatment and making effective strategies for other factors which aid in the disease like: life style and a check on other complex diseases which comes altogether with CKD [17]. Since each population set is different from one another, the scores of risk for a disease differ from ancestry to ancestry. To check differences in PRS in different ancestry’s present study is conducted to find out the differences in African American, South Asians and South East Asians. Polygenic risk score was calculated on the data derived from MatthisWuttke*et. al*. 2019[18].

### Methodology

Data collection: 308 common SNPs of 307 genes associated with eGFRacross different ancestries (SAS, EAS, AA and All ancestries combined) were used for calculating PRS. SNP data was downloaded from [18], PRS was calculated on the basis of effective size of the variants associated with eGFR.(Supplementary data)

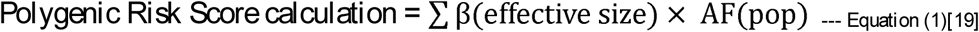

Equation (1) polygenic risk score was calculated by coalesces of effective allele of SNP multiplied with affected allele frequency of any population or of an individual. The score calculated is then normalized by multiplying the β with risk allele dosage (*i*.*e* 2,) and subtracting the population score. All of these normalized values are summarized to get an overall score. The Z score is calculated by using the normalized score divided by summarized population score.

For the calculation of PRS the requirements are: a list of GWAS-significant SNPs, their frequency, effect-size and effect-alleles. This makes it possible to implement the calculation systematically for many diseases and traits. The data visualization was performed using different R packages.

### Results and Discussion

To visualize the Z score values calculated (Table 1), box plot were made for SAS, EAS, AA and all ancestries combined. To have idea about independent risk factor among ancestries and when they are merged what is the effect of polygenic score visualization was done using box plot (Figure 1) : The PRS observed we plotted against the beta calculated for all three ancestries, interestingly, the R^2^ observe showed there is strong correlation between the beta values and PRS (Figure 2).

**Table 1:** Box plot statistics calculated on the basis of PRS for four population sets SAS (South Asians), EAS(East Asians), AA(African American) and all ancestries combined (SAS, EAS, AA).

|  | **All ancestries combined** | **SAS** | **EAS** | **AA** |
| --- | --- | --- | --- | --- |
| Upper Whisker | 6.44 | 2.35 | 1.01 | 1.91 |
| 3<sup>rd</sup> quartile | 2.17 | 0.64 | 0.25 | 0.59 |
| Median | 1.42 | 0.04 | 0.00 | 0.05 |
| 1<sup>st</sup> quartile | -2.01 | -0.56 | -0.26 | -0.46 |
| Lower Whisker | -7.91 | -2.31 | -0.99 | -1.82 |

**Figure 1:**
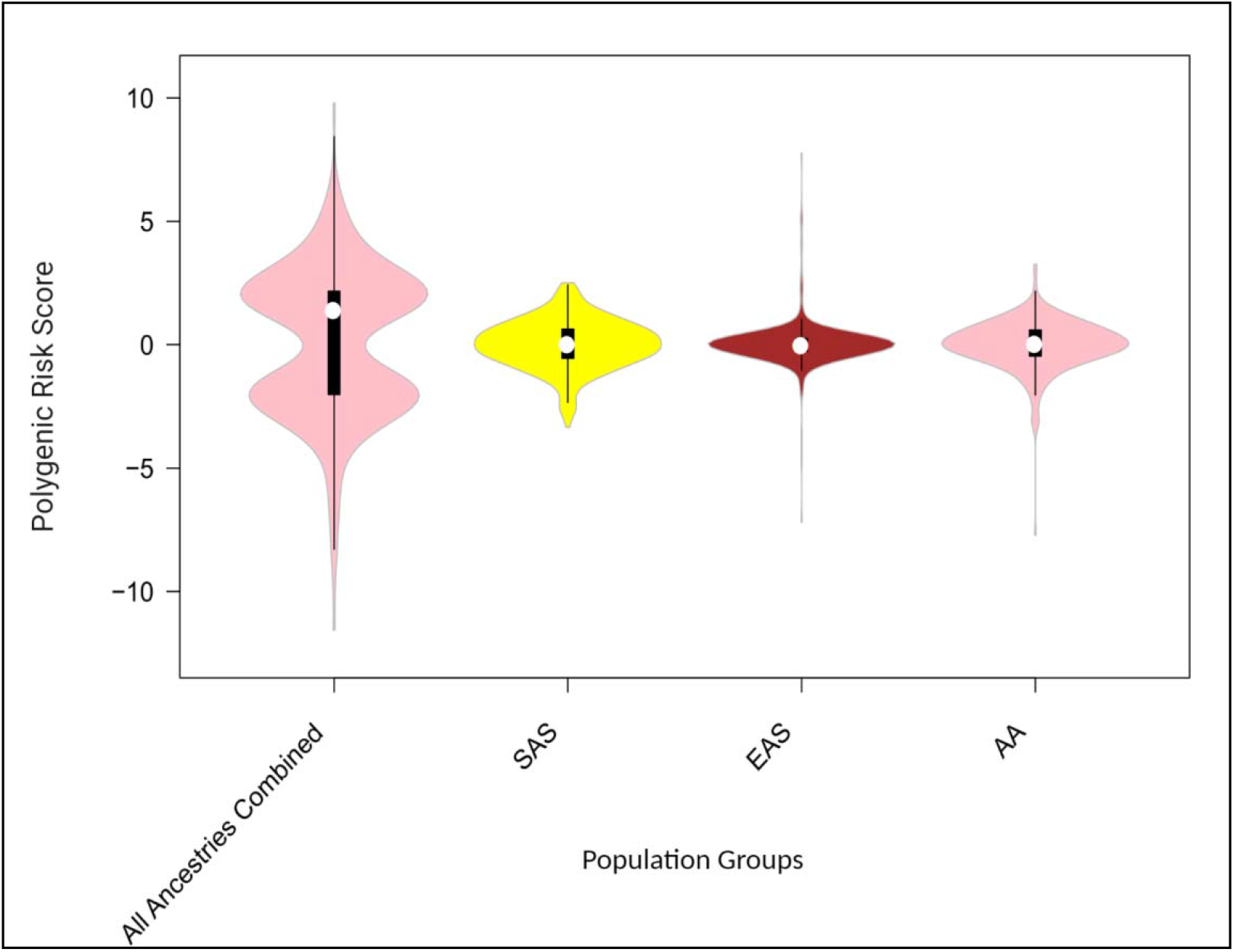
Box plot of Z score values calculated using data for CKD for SAS (South Asians), EAS (East Asians), AA (African Americans) and All ancestries combined (SAS, EAS and AA). The plot signifies that when population groups analyze independently for CKD, the risk factor of the ethnic population group is different and when the population groups of different ancestries are mixed together the risk factor increases and results are biased. This indicates that for the genetic studies individual ethnic groups are needed to be studied to find out the prevalence of the disease in the population group.

**Figure 2:**
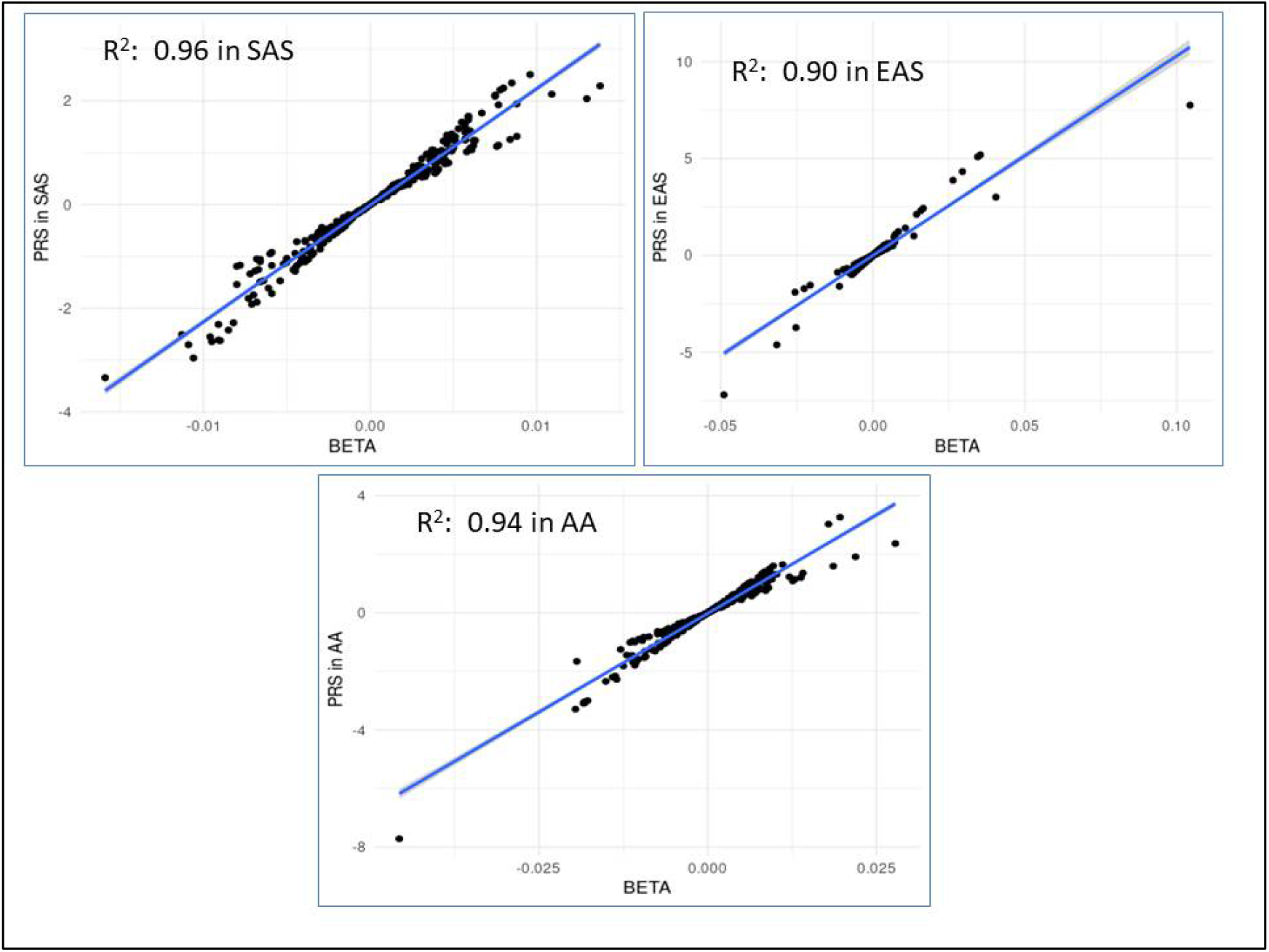
Pearson Correlation of PRS in SAS, EAS and AA showed the risk score obtained from all the 308 SNPs are showing strong correlation with the beta values (effective size)

It is seen that the average risk affinity of the population group with different ancestries is less when they are treated as an individual group, but the tendency of the risk increases when all the population sets are mixed together. This highlights that for genetic studies each population group should be studied independently as per their ethnicity and their medication should be according to their own genetic inferences. This will decrease the biasness in the results which is achieved while merging the samples of different ancestries and will aid in better pharmacogenomics [20]. As seen when the individual population were calculated for risk score SAS were at higher risk for CKD whereas EAS and AA were at little lower risk when compared with SAS and when all the three ancestries were merged, the risk was higher than individual population scores. It is clear that when population groups are pooled the population signatures remains hidden and hence diluting the genetic risk/protection of the population for a particular disease and genetic markers. Hence individual population groups with same ancestry should be targeted for such studies.

Our results are in accordance with the studies conducted [17, 21-24], summarizing that PRS cannot be derived from other ancestry as there can be many differences among the ethnicities in terms of their Linkage Disequilibrium, differences in allele frequencies (variant which is causing risk in one ethnicity might be giving protection to other ethnic group), as a result the PRS would greatly vary as the genetic architecture among ethnic groups varies [17, 25].

GWAS studies are done extensively with respect to different diseases, which is helping massively moving towards personalized medicines, but what we need to consider while conducting such studies is that while framing the study it should be considered that individuals with the same ancestry should be targeted to strengthen the maximum chances of associations measured are related to the targeted disease and not getting diluted or giving increased risk towards disease [26].

Larger sample sets are required for PRS [27] to find out the severity of the disease in particular ethnic group for which GWAS are quite expensive an alternate to it can be small case control studies which are cost effective compared to GWAS can be conducted, it should be practiced to make individual data available online this will help in testing the models when sample sets are merged using small scale local datasets which will serve as a good hold for powerful statistical analysis [28]. The limitation with PRS is the poor performance of it in other than European population due to lack of data from other ancestries [29]. Therefore it is necessary to genotype, sequence and to do case/control studies for rare variants, complex haplotypes, gene-gene interactions for the detection and replication of novel pharmacogenetic loci enhancing the clinicians towards the personalized medicine for all the ethnic groups [20]. This can be achieved by adding local candidate gene association study as well as case control study of that local cohort if in any case GWAS study(ies) are not available. Such studies if conducted will help in knowing the local markers affecting the population groups as the development and outcome of CKD are a brunt of etiological range which is deeply swayed by local risk factors, differences on the basis of genetics, social and demographic changes. Such database if made will aid not only in clinical care but will also help in reducing the disease parameters such as PRS.

## Data Availability

the data used in the manuscript are submitted as supplementary data along with the MS.

## Data Availability

The data used in the present work is contained in the manuscript.

## Author Contributions

IS and VS designed and conceived the study, LG and VS analyzed the data, IS, GR and AS helped in study design, AS critically reviewed the MS.

## Acknowledgement

All the authors acknowledge Mr. Adireddi Govind Rao for providing the computational facilities for the study.

